# Developing Functional Network Connectivity of the Dorsal Anterior Cingulate Cortex Mediates Externalizing Psychopathology in Adolescents with Child Neglect

**DOI:** 10.1101/2021.01.29.21250544

**Authors:** Sarita Silveira, Simone Boney, Susan F. Tapert, Jyoti Mishra

## Abstract

Childhood adversity in form of child abuse and neglect is associated with elevated risk for psychopathologies. We investigated whether development of functional brain networks important for executive function (EF) could serve as potential mediators of this association. We analyzed data of 475 adolescents, a subsample of the multisite longitudinal NCANDA (National Consortium on Alcohol and Neurodevelopment in Adolescence) cohort with completed measures of childhood trauma, resting-state functional brain connectivity data, and internalizing/externalizing psychopathological syndrome data at baseline and follow-up years 1-4. Using parallel process latent growth models, we found that childhood adversity was associated with increased risk for externalizing/internalizing behaviors. We specifically investigated whether functional connectivity of the dorsal anterior cingulate cortex (dACC) to brain regions within the cingulo-opercular (CO) network, a well-known EF network that underlies control of attention and self-regulation, mediates the association between adversity and psychopathological behaviors. We found that childhood adversity, specifically neglect was negatively associated with functional connectivity of the dACC within the CO network, and that this connectivity mediated the association between child neglect and externalizing behaviors. Our study advances a mechanistic understanding of how childhood adversity may impact the development of psychopathology, highlighting the relevance of dACC functional networks particularly for externalizing psychopathology.

## 1. Introduction

Childhood adversity is associated with elevated risk for psychopathologies, including all commonly occurring forms of internalizing and externalizing behaviors in youth (1,2). Research has examined the association between childhood adversity and mental illness in adulthood, but the developmental trajectory of this association in adolescence is still unknown. Adolescence is a critical time period for the emergence of psychopathologies, and attempts have been made to relate this phenomenon to simultaneous changes in neurobiological structure and function and its cognitive and affective correlates (3,4). In this regard, a pivotal role is ascribed to the development of executive function (EF), an overarching human cognitive ability to flexibly select, maintain, and act upon goal-relevant information, while also suppressing irrelevant distractions (5–7). EF, thus, encompasses fundamental cognitive functions such as sustained attention, conflict processing, and self-regulation. Impairments in EF are a transdiagnostic factor determining mental disorders (8). Impaired EF has also been suggested as a potential mechanism to explain the impact of childhood adversity on the development of psychopathologies, which include maladaptive cognitive and behavioral responses to distress based on emotion regulation deficits (1).

EF starts to develop during childhood, yet it improves and refines markedly during adolescence (7,9,10), highlighting the relevance of this period in transitioning to healthy vs. impaired neurocognition. The development of EF during adolescence critically depends on brain maturation, including synaptic pruning (11) and myelination (12), which enhance connectivity within and between brain systems (13). Particularly the dorsal anterior cingulate cortex (dACC) is an important hub region for these processes (14,15). Functionally, the dACC frequently co-activates with the bilateral anterior insula, with which it forms the core of a cingulo-opercular (CO) network in adults (16–18). Functional networks are characterized by dense internal connectivity, and thus network organization of the brain is defined by the network affiliation of each region of the connectome (19). Indeed, brain network function that correlates with EF has been grouped into two networks, the CO and the fronto-parietal (FP) network (16). In task related brain states, while the FP network has been ascribed to the initiation and adjustment of moment-to-moment task control, the CO network provides sustained set-maintenance over the entire task duration. The distinct groupings of these two networks have not only been found during the performance of cognitive tasks, but also during resting state (20). Additionally, age-related neurodevelopmental changes have been observed in these functional network organizations (14,21–23). During childhood, the CO network is not functionally distinct, but is part of a larger set of regions. Adolescence marks the period during which the key dACC region segregates from its FP connections to form the CO network (24). Particularly, increased CO network integration during adolescence has also been shown to moderate the effect of age in the transition to adult-level EF (25). Moreover, in our recent, albeit small sample research in adolescents with childhood adversity, we have found that reduced strength of maturing functional connectivity of the dACC in the CO network is associated with greater child neglect severity (26).

A critical distinction in developmental psychopathology is manifested in patterns of disordered self-control, which relate to internalizing or externalizing problems. These behavioral patterns may occur as early as childhood and pave a developmental pathway to negative social and behavioral outcomes (27–29). While internalizing behavior appears in the form of withdrawal, anxiety and depression, externalizing behaviors relate to hyperactivity, aggression and destructive behavior. Neurodevelopmental mechanisms support two distinct forms of developmental learning that result in the differential problem behaviors (15). Internalizing is supported by an internal self-regulation system that is relevant for attention control under aversive challenges. Externalizing is based on high responsiveness to environmental influences that are appetitive and motivate approach behavior. Traumatic experiences during childhood have been shown to cause adaptive and enduring psychobiological responses that shape these developmental learning processes (30). More consistently reported brain changes in the context of adversity, which have been linked to the development of psychopathology, include structural changes of the hippocampus and anterior cingulate cortex (ACC), amygdala volume and reactivity, as well as a decreased hemispheric integrity of the corpus callosum; however, changes were found to vary with regards to the type of maltreatment that was experienced (31–33). Very few longitudinal studies have investigated a link between childhood adversities, functional brain connectivity and child/adolescent psychopathologies (34,35). These studies highlight the role of functional connectivity in predefined regions corresponding to impulse control and emotion processing, as distinctly relevant to internalizing and externalizing behaviors. However, while a meta-analysis in adults highlights the overall relevance of dACC / anterior insula based network integrity across various psychiatric diagnoses, which is consistent with transdiagnostic deficits in EF (36), no study to date has investigated dACC connectivity as a mediator of childhood adversity and developmental psychopathology.

On the one hand, impaired EF and associated structure and function of brain circuits have been argued as common mechanisms underlying mental illness sequelae in individuals who have suffered childhood adversities. Yet, emerging evidence indicates that these alterations in EF development only occur following specific forms of childhood adversity (37–39). A recent conceptual framework distinguishes adversities in two main dimensions, namely threat and deprivation, with unique emotional, cognitive, and neurobiological developmental pathways relevant to the emergence of psychopathology (40). Child abuse is associated with experiences of violence or risk of violence, and is thus categorized under the dimension of threat. Child neglect involves a lack of parental care and constitutes a form of deprivation of social and emotional inputs. Associations of threat or deprivation with psychopathology are suggested to be mediated by alterations in pruning and potentiation of synapses during neurodevelopment (40). Experiences of deprivation in particular have been found linked to impairments in EF, thereby increasing the risk for psychopathology (38,41). A recent longitudinal study provides evidence that, while both early life deprivation and threat increase risk for internalizing and externalizing psychopathology in adolescents, only the link between childhood deprivation and developing externalizing behaviors is mediated by impaired cognitive abilities (42). In our recent cross-sectional research on a subsample of the current cohort of adolescents, we identified dACC functional connectivity among that of other regions, to mediate the association between childhood adversity and EF self-reports (43). This study investigated the association between childhood adversity, in the forms of child abuse and child neglect, and developing functional brain connectivity underlying EF. At the same time, we investigated the link between the brain network connectivity associated with adversity and the development of internalizing and externalizing behaviors. Specifically, we investigated how child abuse and neglect associate with functional connectivity of the dACC in the CO network, and whether these neurofunctional correlates mediate the development of externalizing and internalizing psychopathologies.

## 2. Materials and Methods

### Sample

We analyzed data of 475 adolescents (mean age 16.97, age range 12-22 years; 52.4% female), a subsample of the 831 multisite NCANDA (National Consortium on Alcohol & Neurodevelopment in Adolescence) cohort. NCANDA uses an accelerated longitudinal design to sample subjects from a broad span of baseline ages (44,45). This allows to address development across larger age ranges than traditional cohort designs. The subsample for the current research was selected according to existing reports on all measurements that are relevant to our hypotheses: (a) self-report on childhood adversity in one of the years of assessment, (b) resting-state functional magnetic resonance imaging (rs-fMRI) data in at least two years of assessment, (c) parent-report for adolescents under age 18 or self-report for those over age 18 on psychopathologies in at least two years of assessment. Psychopathological syndrome data was missing in 9 cases at follow-up year 1, in 14 cases at follow-up year 2, in 40 cases at follow-up year 3 and in 109 cases (23%) at follow-up year 4. Rs-fMRI data was missing in 57 cases at study baseline, in 25 cases at follow-up year 1, in 36 cases at follow-up year 2, in 62 cases at follow-up year 3 and in 133 cases (28%) at follow-up year 4.

The study was approved by the institutional review board of each NCANDA site. All subjects underwent informed assent or consent annually. When under age 18, the participants’ legal guardians provided written informed consent to participate and the youth provided written informed assent. Participants over age 18 provided written informed consent themselves.

### Assessments

#### Childhood adversity

Childhood adversity was assessed using the 28-items brief screening version of the Childhood Trauma Questionnaire, short form (CTQ-SF; (46)). The CTQ-SF is a self-administered retrospective inventory consisting of five categories of child neglect and abuse. Each category entails five items that are rated on a 5-point Likert scale ranging from never true (= 1) to very often true (= 5). For descriptive purposes, subscale scores were summed and those with low to moderate adversity were identified using recommended cut-off scores of subscale sums (Bernstein & Fink, 1998; emotional abuse ≥ 9, physical abuse ≥ 8, sexual abuse ≥ 6, emotional neglect ≥ 10, physical neglect ≥ 8; Table 1). Subscale scores were averaged to inform composite scores for child neglect (i.e., emotional neglect, physical neglect; sample mean = 1.55 ± .60; range = 1-5) and abuse (i.e., emotional abuse, physical abuse, sexual abuse; sample mean = 1.21 ± .33; range = 1-5).

**Table 1.** Samples descriptives (mean ± standard deviation) of time-invariant covariates at baseline and time-varying neurofunction and psychopathology at baseline and at follow-up after one (Y1), two (Y2), three (Y3) and four (Y4) years. SES = socioeconomic status, AUD = alcohol use disorder, dACC = dorsal anterior cingulate cortex, CO = cingulo-opercular network.

| Table 1. Study Sample |  | Baseline |  |  |  |  |
| --- | --- | --- | --- | --- | --- | --- |
| Age |  | 16.97 ± 2.31 |  |  |  |  |
| Sex | Male | 47.6% |  |  |  |  |
|  | Female | 52.4% |  |  |  |  |
| Ethnicity | Caucasian | 73.3% |  |  |  |  |
|  | African American | 13.9% |  |  |  |  |
|  | Asian | 6.3% |  |  |  |  |
|  | Mixed or other | 6.5% |  |  |  |  |
| SES |  | 16.80 ± 2.35 |  |  |  |  |
| Exceeds status |  | 17.1% |  |  |  |  |
| AUD family history |  | .18 ± .38 |  |  |  |  |
| MRI scanner type | Siemens | 38.9% |  |  |  |  |
|  | GE | 61.1% |  |  |  |  |
| Childhood adversity | Neglect | 1.55 ± .60 |  |  |  |  |
|  | Abuse | 1.21 ± .33 |  |  |  |  |
|  | ≥ low to moderate: |  |  |  |  |  |
|  | Emotional abuse | 17.7% |  |  |  |  |
|  | Physical abuse | 16.2% |  |  |  |  |
|  | Sexual abuse | 5.1% |  |  |  |  |
|  | Emotional neglect | 34.7% |  |  |  |  |
|  | Physical neglect | 24.6% |  |  |  |  |
| Longitudinal Measures |  | Baseline | Y1 | Y2 | Y3 | Y4 |
| dACC-CO connectivity |  | .29 ± .13<br>(n=418) | .23 ± .13<br>(n=450) | .28 ± .14<br>(n=439) | .27 ± .14<br>(n=413) | .25 ± .13<br>(n=342) |
| Externalizing psychopathology (T-scores) |  | 42.73 ± 8.51<br>(n=475) | 43.87 ± 9.04<br>(n=466) | 44.00 ± 9.31<br>(n=461) | 44.78 ± 9.71<br>(n=435) | 45.00 ± 9.68<br>(n=366) |
| Internalizing psychopathology (T-scores) |  | 44.63 ± 9.01<br>(n=475) | 46.02 ± 10.30<br>(n=466) | 45.94 ± 10.91<br>(n=461) | 46.68 ± 10.80<br>(n=435) | 47.58 ± 11.24<br>(n=366) |

#### Psychopathology

Internalizing and externalizing behaviors in adolescents under the age of 18 years were determined using parent-reports on the school age version of the Child Behavior Checklist (CBCL) and, if those were not available, self-reports on the Youth Self Report (YSR) of the Achenbach System of Empirically Based Assessments (ASEBA) were used (47). Subjects older than 18 years completed the ASEBA Adult Self Report (ASR). All three scales consist of 112 statements about the adolescent’s behaviors that are rated on a 3-point Likert scale from 0 (= not true) to 2 (= very true). Scores were summed up into composite scales for internalizing and externalizing behaviors, respectively. The internalizing score is composed of syndrome scales for anxious/depressed and withdrawn-depressed behaviors, and somatic complaints. The score for externalizing behaviors consists of rule-breaking and aggressive behavior syndrome scales. The CBCL provides sex and age normed T-scores. In the current study, we are using normative T-scores for the 12-18 years age range. Higher scores indicate greater degrees of behavioral and emotional problems. T-scores of ≥64 on internalizing and externalizing scales indicate deviant behavior that is clinically relevant.

#### Demographics

Demographic variables included age, gender, ethnicity (Caucasian, African American, Asian or mixed) and socio-economic status (SES). SES was determined by the highest level of education achieved by either parent from 1 (1st grade) to 20 (4+ years of graduate/professional school) (48). Parental income was not used as an indicator of SES due to substantial differences in salaries across the NCANDA study sites (49). As the NCANDA study was designed to inform associations between alcohol use and brain development, youth at risk for heavy drinking were oversampled. Control variables therefore included family history of AUDs and early alcohol use prior to the study. AUD family history density index was calculated for first- and second-degree relatives ([# Positive Parents] + [# Positive Grandparents * 0.5]; range of 0-4) (50). Early alcohol use was defined as a dichotomous variable indicating exceeding age-defined drinking thresholds at baseline, potentially indicating some level of alcohol-related risk (“*exceeds”; n=81)* (51). At baseline, higher frequencies of externalizing problems were seen in *exceeds* (7.9%) than in the other participants (5.8%) (52).

### Imaging Acquisition

We used rs-fMRI data that was assessed at baseline and follow-up years 1-4 for all 475 participants to extract functional brain connectivity measures. Neuroimaging data were acquired on two different 3.0T MRI scanner models in NCANDA. 38.9% of participants were scanned with a Magnetom Trio Trim (Siemens Healthcare, Germany) and 61.1% with a MR750 (GE Healthcare, USA). Scan protocols were consistent between sites and scanners. High-resolution anatomical T1 scans (1.2mm slices thickness, repetition time [TR] = 1900ms, echo time [TE] = 2.92ms, 11° flip angle, 240mm field of view [FoV]) and gradient echo-planar imaging (EPI) scans were collected (5mm slice thickness, 32 slices, TR = 2200ms, TE = 30ms, 79° flip angle, FoV = 240, 240×240 matrix).

### Data analysis

Statistical analyses were conducted using MATLAB (The Mathworks, Inc., Natick, MA) and SPSS 19.0 (IBM Corp., Armonk, NY). Rs-fMRI data were analyzed using the CONN functional connectivity toolbox (https://www.nitrc.org/projects/conn) (53). The first five volumes of the EPI scans were discarded to allow for magnetic saturation effects. Data preprocessing included slice time correction, data realignment to the first functional volume, individual rigid T1-to-EPI co-registration, segmentation and normalization of the co-registered T1 and EPI volumes in the MNI stereotactic space. To minimize noise and residual inter-subject differences, spatial smoothing was applied using an isotropic Gaussian kernel with 8mm full width at half maximum. Physiological artifacts and residual subject movements were removed from the data using regression of six rigid-body motion parameters, which were estimated in the realignment step, and regression of outlier scans, based on scrubbing of bad data points as identified by artifact detection. Subject-level analyses additionally included regression of averaged white matter signals and cerebrospinal fluid. On average, 28.1 ± 6.3 % of volumes were discarded from the analyses. None of the subjects were removed completely during the preprocessing procedure.

We analyzed functional connectivity between predefined regions of interest (ROI-to-ROI), which undergo substantial development during adolescence and are known to subserve executive functioning. We determined the relationship of this functional connectivity to childhood adversity and the extent to which this connectivity mediates a link between adversity and psychopathology. Regions within the CO network were manually created as 12mm radius spheres with peak MNI coordinates from previous literature (24). The dACC (MNI coordinates: −2, 7, 50) was defined as a seed region based on prior evidence that it segregates from its FP connections to form the CO network during typical adolescence (24), as well as our prior results that dACC functional connectivity relates to childhood adversity (26) and mediates the link between adversity and self-reported EF in a cross-sectional sample of adolescents (43). Other regions of the CO include the bilateral anterior insula (left: −36, 17, 3; right: 38, 19, 0), anterior prefrontal cortex (left: −28, 53, 16; right: 28, 51, 25) and thalamus (left: −13, −14, 4; right: 11, −14, 6). Functional network connectivity between dACC and CO-ROIs was calculated by median connectivity weights of blood-oxygen level dependent (BOLD) time-series.

Mediation analyses were conducted using AMOS 26.0 (54). We used parallel process latent growth modeling (LGM) as an application of structural equation models for longitudinal mediation analyses. Missing data on repeated measures was addressed using Bayesian multiple imputation (MI) as implemented in AMOS. To capture the uncertainty in predicting data given the observed information, MI generated 10 different data sets, each containing a set of maximum likelihood estimates for missing values. The conversion criterion was satisfied. The LGMs were then fit to each imputed data set, and model fit indices as well as standardized estimates were pooled across all datasets (55,56). In two LGMs, the median connectivity measures extracted in the previous analyses were used to inform latent random intercept and random linear slope of neurofunctional development within the CO network. Similarly, scores on the ASEBA, were used to model intercept and slope of internalizing and externalizing behaviors respectively. Loadings for the slope factors were set equal to values of the time variable, defined with a value of 0 at study baseline and an incremental increase of 1 for each consecutive year of follow-up assessment. Loadings for the intercepts were set equal to 1. Intercept and slope factors represent functions of means and individual deviations from those means. The variances of intercepts and slopes are residual variances. Equality constraints were included on uncorrelated variance residuals of time-varying indicators across measurement occasions (57). Negative error variance estimates were obtained from rs-fMRI data, i.e., Heywood case (58). In accordance with Van Driel’s method (59), residual slope variance of σ^2^ = .00, 95% CI [-.00, .00] and a critical ratio of -.56 indicates that the improper estimate can be seen as a result of sampling fluctuations rather than specification errors (60). Therefore, residual slope variance of rs-fMRI data was constrained to 0 (61). To investigate longitudinal mediation effects, we modeled parallel processes from CTQ-SF neglect and abuse scores to the intercept of dACC-CO connectivity (= baseline status of connectivity) and then to the slope of externalizing/internalizing behaviors (= change in psychopathology over time), as well as from neglect and abuse scores to the intercept of psychopathology and then to the slope of dACC-CO connectivity. This setup assumes that when modeling an intercept as putative mediator of latent growth, a temporal order between mediator and outcome can be presumed (62). We additionally tested whether child neglect and abuse associations with psychopathology, and mediating effects can be traced to functional connectivity between the core CO nodes, the dACC and bilateral anterior insula. Additionally, growth curves of internalizing and externalizing behaviors as well as resting-state functional connectivity were plotted (see supplement S1).

Age, sex, ethnicity, SES, *exceeds* status, AUD family history, and MRI scanner type were included as covariates. Thereby, ethnicity as categorical variable was dummy coded into several dichotomous variables, i.e. Caucasian, African American, Asian, mixed or other group membership. Even though age was assessed at each data collection time point, it was treated as a time-invariant covariate at study baseline. There was missing data on SES scores in 8.8% of cases. These missing values were imputed by using the group mean (63). All covariates except for scanner type were modeled with paths on latent variables (i.e., intercepts and slopes) of both functional connectivity measures and internalizing/externalizing scores. Covariance between sample demographics, abuse and neglect scores were included in the model. Due to non-normality of CTQ-SF and CBCL scores, we performed bootstrapping of maximum likelihood estimation, using 1000 bootstrap samples and a Boot factor of 1. We report bias-corrected confidence intervals of 95%. *χ*^*2*^ statistics, Comparative Fit Index (CFI) and Root Mean Square Error of Approximation (RMSEA) were used as indicators of SEM fit.

## 3. Results

### Covariance of childhood adversity and sample demographics

We found a significant positive covariance of child abuse and neglect scores (*β* = .07, 95% CI [.05, .11], *p* = .001). Further, both childhood adversity scales, i.e., neglect and abuse, significantly covaried with age (neglect: *β* = .14, 95% CI [.01, .26], *p* = .034; abuse: *β* = .01, 95% CI [.03, .17], *p* = .006), indicating that the reported severity of adverse childhood experiences is higher in older adolescents. Besides, SES negatively covaried with both neglect (*β* = -.19, 95% CI [-.32, -.07], *p* = .002) and abuse scores (*β* = -.11, 95% CI [-.19, -.04], *p* = .006). Covariates related to alcohol consumption in relatives and adolescents themselves showed a consistent positive association with exposure to childhood adversity. AUD family history was thereby related to both child neglect (*β* = .03, 95% CI [.01, .05], *p* = .008) and abuse (*β* = .03, 95% CI [.01, .05], *p* = .002), and *exceeds* status was related to child abuse (*β* = .02, 95% CI [.00, .03], *p* = .015).

### Externalizing behavior

The LGM for externalizing behavior had a good overall fit pooled across all 10 multiple imputed datasets with *χ*^*2*^ = 343.38, *p* < .000, CFI = .86 and RMSEA = .07, 95% CI [.06, .08]. Both child neglect and abuse were associated with higher scores on the latent intercept of externalizing scores (Table 2). Besides, we found a negative association between neglect scores and median functional connectivity of the dACC node to regions of the CO network. There was no direct association of neglect or abuse severity and slopes, i.e., a change in functional connectivity or externalizing symptoms over time. Yet, we found that lower functional dACC-CO network connectivity was associated with an increased slope of externalizing behaviors; thereby, mediating the link between child neglect and the development of externalizing behavior. Notably, the reverse pathway of adversity-associated externalizing symptoms mediating the development of functional connectivity was not significant.

**Figure 1.**
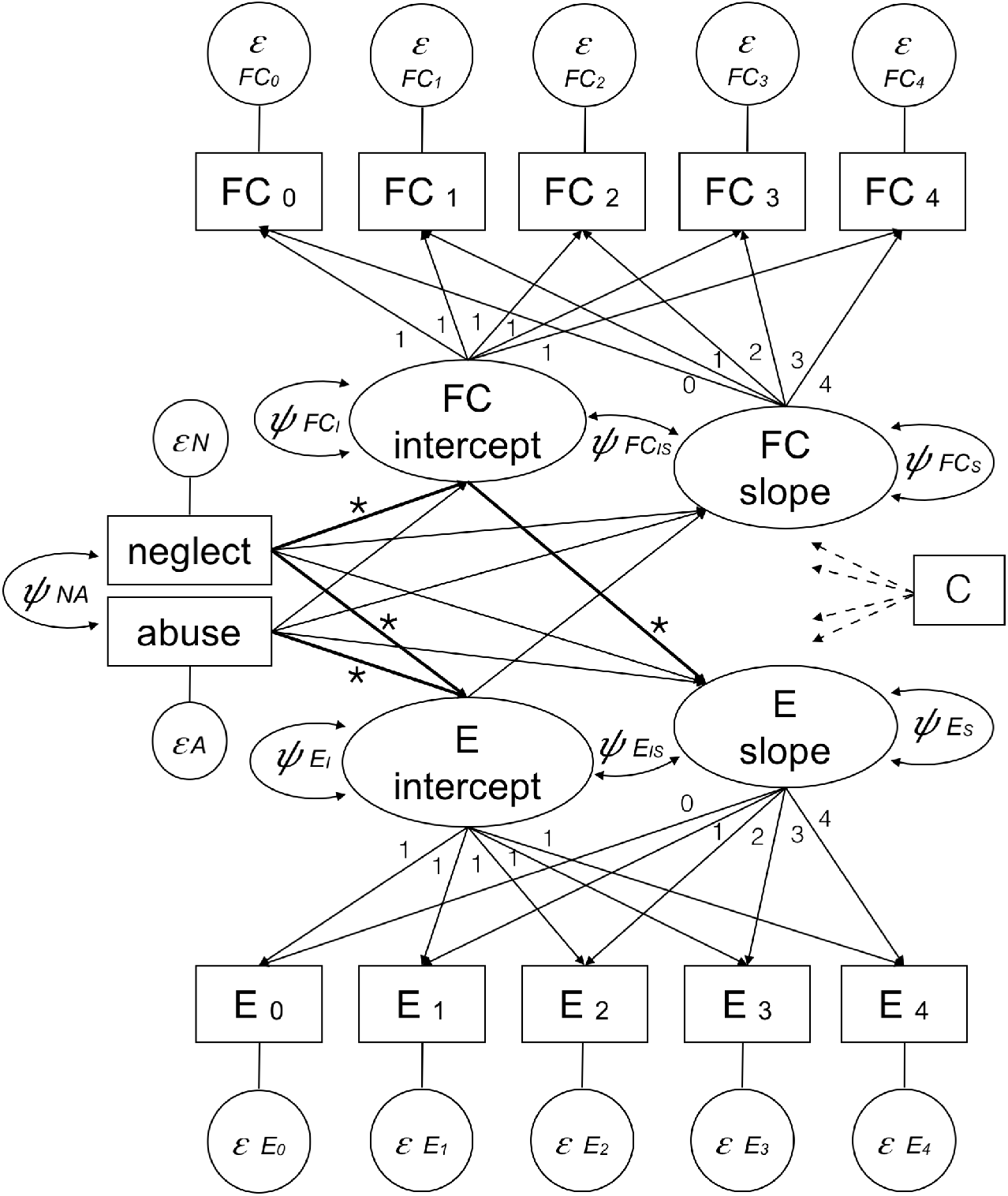
Parallel process latent growth model. *ε* = error term, *ψ* = variance and covariance parameters of latent variables, FC = functional connectivity, E = externalizing behavior, C = covariates (i.e., age, gender, ethnicity, socio-economic status, exceeds status, family history of alcohol use disorder), * = significant paths.

**Table 2.**
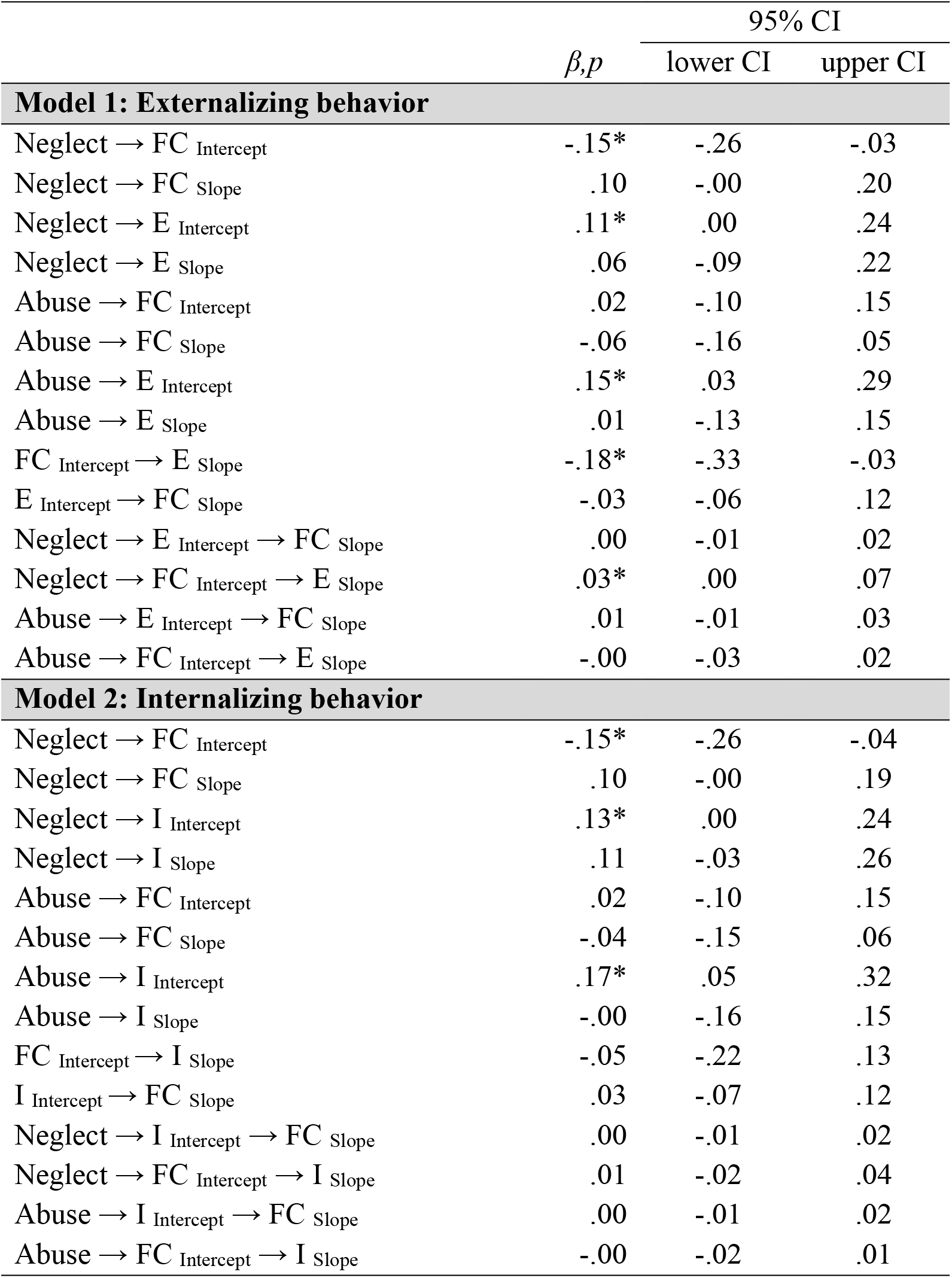
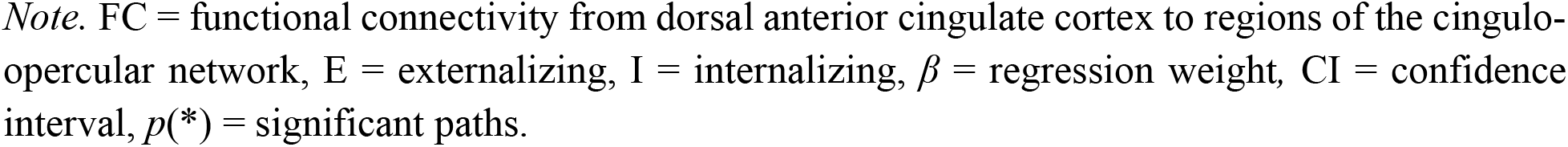
Paths of parallel process latent growth models to link childhood adversity to functional network connectivity and (1) externalizing and (2) internalizing behavior.

| | $\beta, p$ | 95% CI | |
| --- | --- | --- | --- |
|  |  | lower CI | upper CI |
| **Model 1: Externalizing behavior** |  |  |  |
| Neglect → FC Intercept | -.15* | -.26 | -.03 |
| Neglect → FC Slope | .10 | -.00 | .20 |
| Neglect → E Intercept | .11* | .00 | .24 |
| Neglect → E Slope | .06 | -.09 | .22 |
| Abuse → FC Intercept | .02 | -.10 | .15 |
| Abuse → FC Slope | -.06 | -.16 | .05 |
| Abuse → E Intercept | .15* | .03 | .29 |
| Abuse → E Slope | .01 | -.13 | .15 |
| FC Intercept → E Slope | -.18* | -.33 | -.03 |
| E Intercept → FC Slope | -.03 | -.06 | .12 |
| Neglect → E Intercept → FC Slope | .00 | -.01 | .02 |
| Neglect → FC Intercept → E Slope | .03* | .00 | .07 |
| Abuse → E Intercept → FC Slope | .01 | -.01 | .03 |
| Abuse → FC Intercept → E Slope | -.00 | -.03 | .02 |
| **Model 2: Internalizing behavior** |  |  |  |
| Neglect → FC Intercept | -.15* | -.26 | -.04 |
| Neglect → FC Slope | .10 | -.00 | .19 |
| Neglect → I Intercept | .13* | .00 | .24 |
| Neglect → I Slope | .11 | -.03 | .26 |
| Abuse → FC Intercept | .02 | -.10 | .15 |
| Abuse → FC Slope | -.04 | -.15 | .06 |
| Abuse → I Intercept | .17* | .05 | .32 |
| Abuse → I Slope | -.00 | -.16 | .15 |
| FC Intercept → I Slope | -.05 | -.22 | .13 |
| I Intercept → FC Slope | .03 | -.07 | .12 |
| Neglect → I Intercept → FC Slope | .00 | -.01 | .02 |
| Neglect → FC Intercept → I Slope | .01 | -.02 | .04 |
| Abuse → I Intercept → FC Slope | .00 | -.01 | .02 |
| Abuse → FC Intercept → I Slope | -.00 | -.02 | .01 |

*Note.* FC = functional connectivity from dorsal anterior cingulate cortex to regions of the cingulo-opercular network, E = externalizing, I = internalizing, $\beta$ = regression weight, CI = confidence interval, $p(*)$ = significant paths.

Additionally, there was a significant association of *exceeds* status (*β =* .19, 95% CI [.07, .30]) as well as SES (*β = -*.10, 95% CI [-.20, -.01]) with the latent intercept of externalizing behaviors. Scanner type (*β = -*.14, 95% CI [-.25, -.05]) and age (*β =* .14, 95% CI [.02, .26]) were also significantly associated with the intercept of functional connectivity. AUD family history was found to be associated with latent intercepts of externalizing scores (*β* = .16, 95% CI [.06, .27]) and functional connectivity (*β =* -.13, 95% CI [-.25, .01]), and with the latent slope of functional connectivity (*β = -*.12, 95% CI [-.21, -.02]). Results were not exclusively driven by dACC-anterior insula functional connectivity (see supplement S2).

### Internalizing behavior

The internalizing behavior model had an overall acceptable model fit with *χ*^*2*^ = 391.50, *p* < .001, CFI = .89 and RMSEA = .07, 95% CI [.06, .07]. Again, there was a significant negative association of neglect scores with the latent intercept of dACC-CO network connectivity and a positive association of both neglect and abuse with the latent intercept of internalizing behaviors. There was no direct or indirect link between child neglect or abuse severity and dACC-CO network neurodevelopment or the development of internalizing behaviors over time.

The association of scanner type (*β =* -.14, 95% CI [-.25, -.04]) and age (*β =* .14, 95% CI [.02, .14]) with the latent intercept of functional network connectivity, and of AUD family history with the latent intercept (*β = -*.13, 95% CI [-.25, -.01]) and slope (*β = -*.11, 95% CI [-.21, -.02]) of functional connectivity and with the latent intercept of internalizing behaviors (*β =* .12, 95% CI [.00, .23]) were found to be analogous to the externalizing behavior model. We additionally found an association of age with both latent intercept (*β =* .12, 95% CI [.00, .23]) and slope (*β =* -.18, 95% CI [-.30, -.06]) of internalizing symptoms.

## 4. Discussion

We used the accelerated longitudinal NCANDA dataset to investigate developmental trajectories linking childhood adversity, functional brain connectivity, and psychopathology in adolescence. Regarding the prevalence of adverse childhood experiences, our sample is comparable to previously reported lifetime prevalence metrics in the general population (64). The occurrence of child neglect, particularly emotional neglect, is thereby much higher than the occurrence of child abuse. We found a significant association of child neglect with functional connectivity from the dACC to other regions of the CO network. The dACC plays a crucial role in the development of EF (7,14), particularly as a core region of the CO network that is undergoing segregation and maturation during adolescence (16,21–25). While there was no direct association of childhood adversity with developmental growth in brain connectivity or psychopathology, we found that the link between adversity and the development of externalizing behaviors is mediated by functional dACC network connectivity. Thus, our findings highlight that this mediating effect of dACC-CO connectivity is specific to child neglect. While childhood adversity is generally associated with higher risk of mental illness, different pathways of developmental psychopathology including alterations in structural and functional neurodevelopment have been suggested to relate to the specific types of adversity experiences (39,40). Our result is in accordance with previous findings, which indicate that particularly neglect as a form of deprivation, is linked to psychopathogenesis via impaired neural functions underlying EF (42,65,66).

We further found positive main effects of child neglect and abuse severity on both externalizing and internalizing behaviors in youth. This is in line with the notion that childhood adversity is a risk factor for psychopathology and its onset in adolescence (1,2). However, no mediation by dACC-CO connectivity was found for the development of internalizing behaviors. Previous research suggests that dysregulated self-control can predict both internalizing and externalizing behaviors in children and adolescents (67) and mediate the association of childhood adversity with both internalizing and externalizing behaviors (1). Studies in adults further highlight that impaired structural as well as functional dACC / anterior insula based network integrity is a consistent and transdiagnostic factor in mental illness (36). Yet, in this large sample, we found dACC-CO network connectivity to specifically mediate the link between child neglect and the development of externalizing behavior. This result also dovetails with our prior research showing that interventions that strengthen dACC-CO functional connectivity in neglected adolescents, primarily resolve hyperactivity that is core to externalizing behaviors (22).

Interestingly, recent longitudinal studies highlight a similar link between childhood adversities and EF or EF related functional connectivity that can be related to externalizing, but not internalizing psychopathologies (34,42). The distinction in developmental trajectories of externalizing and internalizing behaviors may be due to the nature of impaired self-control. While self-regulation of internalizing behavior is contextualized in the presence of aversive stimuli, self-regulation of externalizing behavior is relevant in the context of appetitive stimuli (15). Our results indicate a possible mechanistic pathway for how EF impairments following child neglect particularly affect the phenotypic expressions of externalizing behavior characterized by a lack of inhibitory control towards environmental stimuli. The CO network shows considerable overlap with the salience network, particularly in the core nodes of dACC and bilateral anterior insula (18,68). Functional coupling between these core regions has been associated with the presentation of salient events as well as with transient attention (17). Thus, impacted functional connectivity from the CO/salience network core nodes may lead to heightened risk for externalizing symptoms due to a lack of self-control in the presence of salient environmental stimuli. However, when testing for a mediating effect specifically of dACC connectivity with the bilateral insula, the results could not be traced to connectivity between these CO core nodes; our results, thus, appear to be a product of overall dACC-CO within-network connectivity.

Additionally, using parallel process latent growth models, we simultaneously tested the reverse relationship between internalizing or externalizing behaviors and the development of functional brain connectivity. Neither internalizing nor externalizing phenotypes of adolescent psychopathology could be linked to developmental trajectories of dACC-CO network connectivity. These results support the notion that functional connectivity patterns can be a useful biomarker relevant to individual differences in behavior, but not vice versa, with implications for neurally-targeted personalized medicine (69). While the majority of functional connectivity based prediction studies have focused on classifying disease status at the time of the fMRI scan, this study contributes to a research gap on individual variations and longitudinal associations with dimensional behavior-based clinical phenotypes (70,71).

NCANDA oversamples youth at higher risk for risky alcohol use patterns. We therefore included AUD risk related covariates in our analyses, i.e., drinking patterns that exceed age-related thresholds and an index for AUD family history. We found high covariance between both these risk factors and childhood adversity, indicating that adolescents who experienced childhood adversity are more likely to have parents or grandparents with AUDs, and that those who experienced child abuse are more likely to drink more alcohol than others of the same age. *Exceeds* status was further associated with higher levels of externalizing behaviors. AUD family history was related to higher levels of both externalizing and internalizing behaviors as well as with lower baseline status and decelerated development of dACC-CO functional network connectivity. This indicates that alcohol use related factors within the family may be important precursors of both baseline status and development of psychopathology in youth. Previous literature provides evidence that particularly children of alcoholics show elevated levels of internalizing and externalizing symptoms in childhood and adolescence (72,73).

One limitation of the current research is a sole focus on dACC resting state functional connections. Due to this specific scope, the current study is not exhaustive in explaining (a) alternative neurofunctional pathways that may be relevant to the development of externalizing psychopathology and (b) potential neurofunctional pathways that may be relevant to the development of internalizing psychopathology. The focus on dACC connectivity in particular is based on a body of research that suggests its formation during adolescence, and its association with EF (14,25) and childhood adversity (32,33). A further limitation of this study is that no EF task is included in the analyses. Interpretation of results are thus based on previous findings that link dACC and CO network connectivity to EF and its development during adolescence. Therefore, no explicit conclusions can be drawn about EF in the current sample. Future studies are needed to investigate a direct link between dACC-CO network connectivity, the development of EF performance and psychopathology.

Taken together, our study advances a mechanistic understanding of how childhood adversity may be linked to the development of psychopathology during adolescence. Notably, it supports the notion of different developmental trajectories dependent on the type of experienced adversity, neglect vs. abuse. Further, it highlights the relevance of dACC based functional networks in mediating a link between child neglect and externalizing behaviors. Results from this study may inform the body of research directed towards neurotherapeutic approaches by identifying specific functional brain connectivity as possible intervention and prevention target.

## Data Availability

Data from the National Consortium on Alcohol and Neurodevelopment in Adolescence (NCANDA) is available upon request. For further information, please visit https://www.niaaa.nih.gov/ncanda-data-distribution-agreement

## Acknowledgements

This research was supported by University of California San Diego (UCSD) lab start-up funds (Mishra) and the German National Academy of Sciences Leopoldina fellowship (Silveira). Reported data was obtained from the multisite National Consortium on Alcohol & Neurodevelopment in Adolescence (NCANDA) study, which is supported by the U.S. National Institute on Alcohol Abuse and Alcoholism with co-funding from the National Institute on Drug Abuse, the National Institute of Mental Health, and the National Institute of Child Health and Human Development grant numbers: AA021697 (Pfefferbaum and Pohl), AA021695 (Brown and Tapert), AA021692 (Tapert), AA021696 (Colrain and Baker), AA021681 (De Bellis), AA021690 (Clark), and AA021691 (Nagel).

## References

1. Heleniak C, Jeness JL, Stoep A Vander, McCauley E, McLaughlin KA. Childhood maltreatment exposure and disruptions in emotion regulation: A transdiagnostic pathway to adolescent internalizing and externalizing psychopathology. Cognit Ther Res. 2016;40(3):394–415.

2. McLaughlin KA, Green JG, Gruber MJ, Sampson NA, Zaslavsky AM, Kessler RC. Childhood Adversities and First Onset of Psychiatric Disorders in a National Sample of US Adolescents. Arch Gen Psychiatry. 2012;69(11):1151–60.

3. Giedd JN, Keshavan M, Paus T. Why do many psychiatric disorders emerge during adolescence? Nat Rev Neurosci. 2008;9(12):947–57.

4. Gogtay N, Giedd JN, Lusk L, Hayashi KM, Greenstein D, Vaituzis AC, et al. Dynamic mapping of human cortical development during childhood through early adulthood. Proc Natl Acad Sci [Internet]. 2004;101(21):8174–9. Available from: http://www.pnas.org/cgi/doi/10.1073/pnas.0402680101

5. Badre D. Defining an Ontology of Cognitive Control Requires Attention to Component Interactions. Top Cogn Sci. 2011;3:217–21.

6. Lenartowicz A, Kalar DJ, Congdon E, Poldrack A. Towards an Ontology of Cognitive Control. Top Cogn Sci. 2010;2:678–92.

7. Luna B, Marek S, Larsen B, Tervo-Clemmens B, Chahal R. An integrative model of the maturation of cognitive control. Annu Rev Neurosci. 2015;38:151–170. PMCID: PMC5661874.

8. McTeague LM, Goodkind MS, Etkin A. Transdiagnostic impairment of cognitive control in mental illness. J Psychiatr Res. 2016;83:37–46.

9. Best JR, Miller PH. A Developmental Perspective on Executive Function. Child Dev. 2010;81(6):1641–60.

10. Crone EA. Executive functions in adolescence: Inferences from brain and behavior. Dev Sci. 2009;12(6):825–30.

11. Petanjek Z, Judaš M, šimic G, Rašin MR, Uylings HBM, Rakic P, et al. Extraordinary neoteny of synaptic spines in the human prefrontal cortex. Proc Natl Acad Sci. 2011;108(32):13281–6.

12. Simmonds D, Hallquist MN, Asato M, Luna B. Developmental Stages and Sex Differences of White Matter and Behavioral Development through Adolescence: A Longitudinal Diffusion Tensor Imaging (DTI) Study. Neuroimage. 2014;15(92):356–68.

13. Luna B, Sweeney JA. The emergence of collaborative brain function: fMRI studies of the development of response inhibition. Ann N Y Acad Sci. 2004;1021:296–309.

14. Ordaz SJ, Foran W, Velanova K, Luna B. Longitudinal Growth Curves of Brain Function Underlying Inhibitory Control through Adolescence. J Neurosci. 2013;33(46):18109-18124. PMCID: PMC3828464.

15. Tucker DONM, Poulsen C, Luu P. Critical periods for the neurodevelopmental processes of externalizing and internalizing. Dev Psychopathol. 2015;27:321–46.

16. Dosenbach NUF, Visscher KM, Palmer ED, Miezin FM, Wenger KK, Kang HC, et al. A core system for the implementation of task sets. Neuron. 2006;50(5):799-812. PMCID: PMC3621133.

17. Han SW, Eaton HP, Marois R. Functional Fractionation of the Cingulo-opercular Network: Alerting Insula and Updating Cingulate. Cereb Cortex. 2019;29(6):2624–38.

18. Power JD, Petersen SE. Control-related systems in the human brain. Curr Opin Neurobiol [Internet]. 2013;23(2):223–8. Available from: http://dx.doi.org/10.1016/j.conb.2012.12.009

19. Newman MEJ. Modularity and community structure in networks. Proc Natl Acad Sci. 2006;103(23):8577–82.

20. Power JD, Cohen AL, Nelson SM, Wig GS, Barnes KA, Church JA, et al. Functional network organization of the human brain. Neuron. 2011;72(4):665-678. PMCID: PMC3222858.

21. Fair DA, Cohen AL, Power JD, Dosenbach NUF, Church JA, Miezin FM, et al. Functional Brain Networks Develop from a ‘“Local to Distributed”’ Organization. PLoS Comput Biol. 2009;5(5):14–23. PMCID: PMC2671306.

22. Dosenbach NUF, Nardos B, Cohen AL, Fair DA, Power JD, Church J a, et al. Prediction of individual brain maturity using fMRI. Science (80-). 2010;329(September):1358–61.

23. Kelly AMC, Di Martino A, Uddin LQ, Shehzad Z, Gee DG, Reiss PT, et al. Development of anterior cingulate functional connectivity from late childhood to early adulthood. Cereb Cortex. 2009;19(3):640–57.

24. Fair DA, Dosenbach NUF, Church JA, Cohen AL, Brahmbhatt S, Miezin FM, et al. Development of distinct control networks through segregation and integration. Proc Natl Acad Sci. 2007;104(33):13507–13512. PMCID: PMC1940033.

25. Marek S, Hwang K, Foran W, Hallquist MN, Luna B. The Contribution of Network Organization and Integration to the Development of Cognitive Control. PLOS Biol. 2015;13(12):1–25.

26. Mishra J, Sagar R, Parveen S, Kumaran S, Modi K, Maric V, et al. Closed-Loop Digital Meditation for Neuro-Cognitive & Behavioral Development in Adolescents with Childhood Neglect. Nat Transl Psychiatry. 2020;10:1–13.

27. King SM, Iacono WG, Mcgue M. Childhood externalizing and internalizing psychopathology in the prediction of early substance use. Addiction. 2004;99:1548–59.

28. Reef J, Diamantopoulou S, Meurs I, Verhulst F, Ende J. Predicting adult emotional and behavioral problems from externalizing problem trajectories in a 24-year longitudinal study. Eur Child Adolesc Psychiatry. 2010;19:577–85.

29. Hofstra MB, Van Der Ende J, Verhulst FC. Child and adolescent problems predict DSM-IV disorders in adulthood: A 14-year follow-up of a Dutch epidemiological sample. J Am Acad Child Adolesc Psychiatry. 2002;41(2):182–9.

30. Baldwin D V. Primitive mechanisms of trauma response: An evolutionary perspective on trauma-related disorders. Neurosci Biobehav Rev [Internet]. 2013;37(8):1549–66. Available from: http://dx.doi.org/10.1016/j.neubiorev.2013.06.004

31. Teicher MH, Samson JA. Childhood maltreatment and psychopathology: A case for ecophenotypic variants as clinically and neurobiologically distinct subtypes. Am J Psychiatry. 2013;170(10):1114–33.

32. Teicher MH, Samson JA, Anderson CM, Ohashi K. The effects of childhood maltreatment on brain structure, function and connectivity. Nat Rev Neurosci [Internet]. 2016;17(10):652-666. PMID: 27640984. Available from: http://dx.doi.org/10.1038/nrn.2016.111

33. Teicher MH, Samson JA. Annual Research Review: Enduring neurobiological effects of childhood abuse and neglect. J Child Psychol Psychiatry Allied Discip. 2016;57(3):241–266. PMCID: PMC4760853.

34. Barch DM, Belden AC, Tillman R, Whalen D, Luby JL. Early Childhood Adverse Experiences, Inferior Frontal Gyrus Connectivity, and the Trajectory of Externalizing Psychopathology. J Am Acad Child Adolesc Psychiatry [Internet]. 2018;57(3):183–90. Available from: https://doi.org/10.1016/j.jaac.2017.12.011

35. Herringa RJ, Burghy CA, Stodola DE, Fox ME, Davidson RJ, Essex MJ. Enhanced prefrontal-amygdala connectivity following childhood adversity as a protective mechanism against internalizing in adolescence. Biol Psychiatry Cogn Neurosci Neuroimaging. 2016;1(4):326–34.

36. Goodkind M, Eickhoff SB, Oathes DJ, Jiang Y, Chang A, Jones-Hagata LB, et al. Identification of a comon neurobiological substrate for mental illness. JAMA Psychiatry. 2015;72(4):305–15.

37. Lambert HK, King KM, Monahan KC, McLaughlin KA. Differential associations of threat and deprivation with emotion regulation and cognitive control in adolescence. Dev Psychopathol. 2017;29:929–40.

38. Sheridan MA, Peverill M, Finn AS, McLaughlin KA. Dimensions of childhood adversity have distinct associations with neural systems underlying executive functioning. Dev Psychopathol. 2017;29:1777–94.

39. Teicher MH, Samson JA, Anderson CM, Ohashi K. The effects of childhood maltreatment on brain structure, function and connectivity. Nat Rev Neurosci [Internet]. 2016;17(10):652–66. Available from: http://dx.doi.org/10.1038/nrn.2016.111

40. Sheridan MA, McLaughlin KA. Neurobiological models of the impact of adversity on education. Vol. 10, Current Opinion in Behavioral Sciences. Elsevier Ltd; 2016. p. 108–13.

41. McLaughlin KA, Sheridan MA, Lambert HK. Childhood adversity and neural development: Deprivation and threat as distinct dimensions of early experience. Vol. 47, Neuroscience and Biobehavioral Reviews. Elsevier Ltd; 2014. p. 578–91.

42. Miller AB, Sheridan MA, Hanson JL, McLaughlin KA, Bates JE, Lansford JE, et al. Dimensions of deprivation and threat, psychopathology, and potential mediators: A multiyear longitudinal analysis. J Abnorm Psychol. 2018 Feb 1;127(2):160–70.

43. Silveira S, Nooner KB, Nagel BJ, Tapert SF, De Bellis MD, Mishra J. Cognitive control networks and related vulnerabilities for alcohol abuse in adolescents with childhood trauma. In: Program No 64504 2018 Neuroscience Meeting Planner. San Diego, CA: Society for Neuroscience; 2018.

44. Duncan SC, Duncan TE, Hops H. Analysis of longitudinal data within accelerated longitudinal designs. Psychol Methods. 1996;1(3):236–48.

45. Miyazaki Y, Raudenbush SW. Tests for linkage of multiple cohorts in an accelerated longitudinal design. Psychol Methods. 2000;5(1):44.

46. Bernstein DP, Stein JA, Newcomb MD, Walker E, Pogge D, Ahluvalia T, et al. Development and validation of a brief screening version of the Childhood Trauma Questionnaire. Child Abus Negl. 2003;27(2):169–190. PMID: 12615092.

47. Achenbach TM, Rescorla L. Manual for the ASEBA Preschool forms and profiles. Burlington, VT: University of Vermont Research Center for Children, Youth, & Families; 2000.

48. Rueden U Von, Gosch A, Rajmil L, Bisegger C, Ravens-Sieberer U. Socioeconomic determinants of health related quality of life in childhood and adolescence: results from a European study. J Epidemiol Community Heal. 2006;60(2):130–5.

49. Sullivan E V, Brumback T, Tapert SF, Fama R, Prouty D, Brown SA, et al. Cognitive, emotion control, and motor performance of adolescents in the NCANDA study: Contributions from alcohol consumption, age, sex, ethnicity, and family history of addiction. Neuropsychology. 2016;30(4):449–73.

50. Stoltenberg SF, Mudd S, Blow FC, Hill EM. Evaluating measures of family history of alcoholism: density versus dichotomy. Addiction. 1998;93(10):1511–20.

51. Pfefferbaum A, Kwon D, Brumback T, Thompson WK, Cummins K, Tapert SF, et al. Altered brain developmental trajectories in adolescents after initiating drinking. Am J Psychiatry. 2017;175(4):370–80.

52. Brown SA, Brumback TY, Tomlinson K, Cummins K, Thompson WK, Nagel BJ, et al. The National Consortium on Alcohol and Neuro-Development in Adolescence (NCANDA): A Multisite Study of Adolescent Development and Substance Use. J Stud Alcohol Drugs. 2015;213:895–908.

53. Whitfield-Gabrieli S, Nieto-Castanon A. Conn: A Functional Connectivity Toolbox for Correlated and Anticorrelated Brain Networks. Brain Connect [Internet]. 2012;2(3):125–41. Available from: http://online.liebertpub.com/doi/abs/10.1089/brain.2012.0073

54. Arbuckle JL. Amos 26.0 user’s guide. Chicago: IBM SPSS; 2019.

55. van Ginkel JR. Standardized Regression Coefficients and Newly Proposed Estimators for R2 in Multiply Imputed Data. Psychometrika. 2020 Mar 1;85(1):185–205.

56. Wu W, Jia F. A New Procedure to Test Mediation With Missing Data Through Nonparametric Bootstrapping and Multiple Imputation. Multivariate Behav Res. 2013 Sep;48(5):663–91.

57. Grimm KJ, Widman KF. Residual structures in latent growth curve modeling. Struct Equ Model. 2010;17(3):424–42.

58. Rindskopf D. Structural equation models: Empirical identification, Heywood cases, and related problems. Sociol Methods Res. 1984;13(1):109–19.

59. Van Driel O. On various causes of improper solutions in maximum likelihood factor analysis. Psychometrika. 1978;43:225–43.

60. Chen F, Bollen KA, Paxton P, Curran PJ, Kirby JB. Improper solutions in structural equation models: Causes, consequences, and strategies. Sociol Methods Res. 2001;29(4):468–508.

61. Dillon WR, Kumar A, Mulani N. Offending Estimates in Covariance Structure Analysis: Comments on the Causes of and Solutions to Heywood Cases. Psychol Bull. 1987;101(1):126–35.

62. von Soest T, Hagtvet KA. Mediation analysis in a latent growth curve modeling framework. Struct Equ Model. 2011;18(2):289–314.

63. Cheung MW. Missing Time-Invariant Covariates in Latent Growth Models Under the Assumption of Missing Completely at Random. Organ Res Methods. 2007;10(4):609–34.

64. Kim H, Wildeman C, Jonson-Reid M, Drake B. Lifetime prevalence of investigating child maltreatment among US children. Am J Public Health. 2017;107(2):274–80.

65. McLaughlin KA, Weissman D, Bitrán D. Childhood Adversity and Neural Development: A Systematic Review. Annu Rev Dev Psychol. 2019;1.

66. Spratt EG, Friedenberg SL, Swenson CC, LaRosa A, De Bellis MD, Macias MM, et al. The effects of early neglect on cognitive, language, and behavioral functioning in childhood. Psychology (Irvine). 2012;3(2):175.

67. Hughes C, Ensor R. Individual differences in growth in executive function across the transition to school predict externalizing and internalizing behaviors and self-perceived academic success at 6 years of age. J Exp Child Psychol. 2011;108(3):663–76.

68. Seeley WW, Menon V, Schatzberg AF, Keller J, Glover GH, Kenna H, et al. Dissociable Intrinsic Connectivity Networks for Salience Processing and Executive Control. J Neurosci. 2007;27(9):2349–56.

69. Finn ES, Constable RT. Individual variations in functional brain connectivity: Implications for personalized approaches to psychiatric illness. Dialogues Clin Neurosci. 2016;18(3):277–87.

70. Rosenberg MD, Casey BJ, Holmes AJ. Prediction complements explanation in understanding the developing brain. Nat Commun [Internet]. 2018;9(1):1–13. Available from: http://dx.doi.org/10.1038/s41467-018-02887-9

71. Silveira S, Shah R, Nooner KB, Nagel BJ, Tapert SF, de Bellis MD, et al. Impact of Childhood Trauma on Executive Function in Adolescence—Mediating Functional Brain Networks and Prediction of High-Risk Drinking. Biol Psychiatry Cogn Neurosci Neuroimaging [Internet]. 2020;5(5):499–509. Available from: https://doi.org/10.1016/j.bpsc.2020.01.011

72. Hussong AM, Wirth RJ, Edwards MC, Curran PJ, Chassin LA, Zucker RA. Externalizing symptoms among children of alcoholic parents: Entry points for an antisocial pathway to alcoholism. J Abnorm Child Psychol. 2007;116(3):529–42.

73. Hussong AM, Cai L, Curran PJ, Flora DB, Chassin L, Zucker RA. Disaggregating the distal, proximal, and time-varying effects of parent alcoholism on children’s internalizing symptoms. J Abnorm Child Psychol. 2008;36(3):335–46.

