## Supplement S1 for "Developing Functional Network Connectivity of the Dorsal Anterior Cingulate Cortex Mediates Externalizing Psychopathology in Adolescents with Child Neglect"

**Supplement S1.** Growth trajectories of internalizing and externalizing psychopathology and resting-state functional connectivity from study baseline (Bl) across all follow-up assessments after one (Y1), two (Y2), three (Y3) and four (Y4) years. Plotted are mean growth (blue), standard deviation (grey) for psychopathology, as well as mean growth (blue) and individual growth curves for functional connectivity.

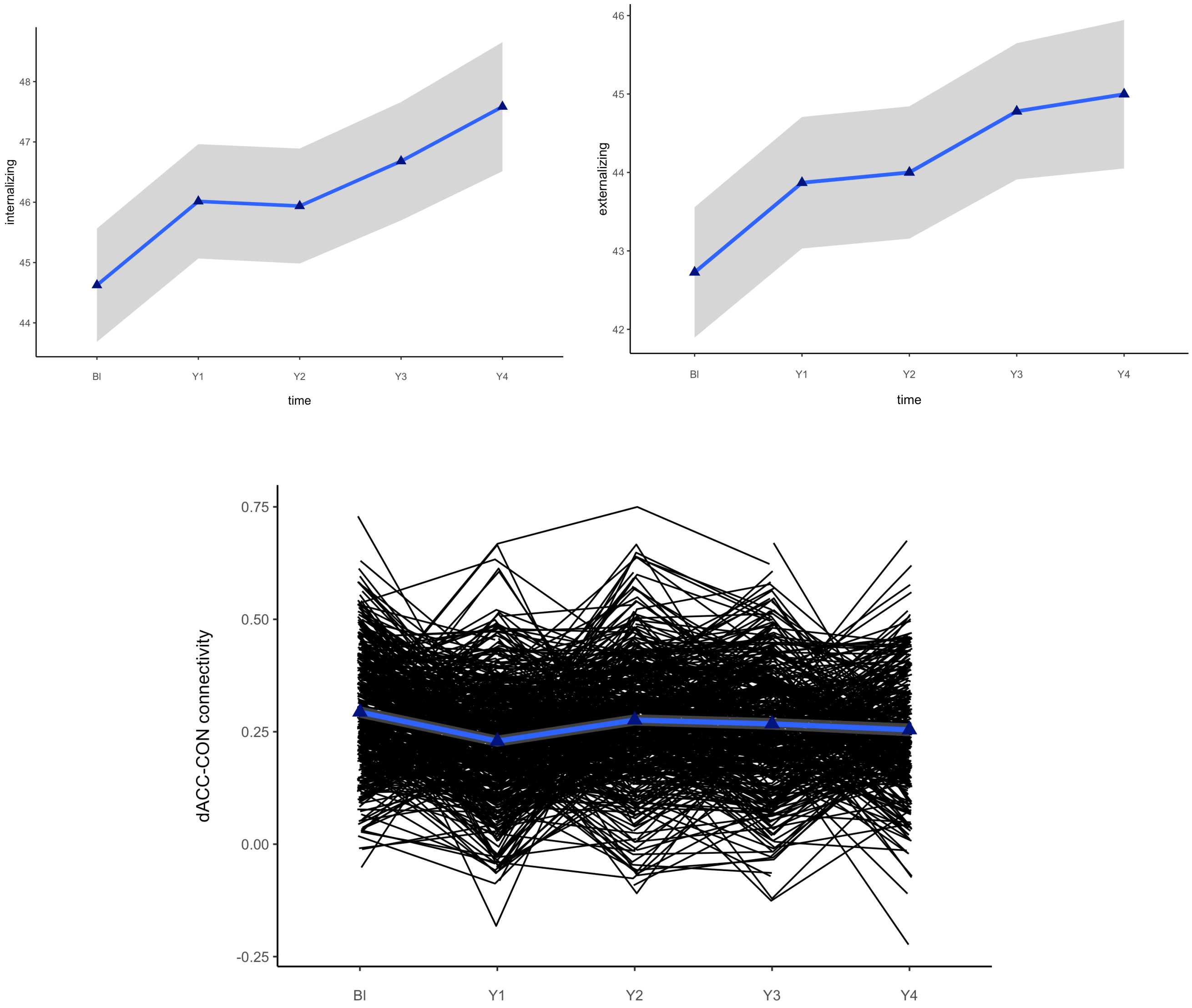
