## Supplement S2 for "Developing Functional Network Connectivity of the Dorsal Anterior Cingulate Cortex Mediates Externalizing Psychopathology in Adolescents with Child Neglect"

**Supplement S2.** Link between child abuse and neglect, functional connectivity from the dorsal anterior cingulate cortex (dACC) to bilateral anterior insula and externalizing/internalizing psychopathology.

To test whether the link between childhood trauma and psychopathology can be mediated by functional connectivity between the CON network's core nodes within the dACC and bilateral anterior insula, we computed two parallel process latent growth models. Both models with longitudinal outcome variables of bilateral dACC-anterior insula functional connectivity and externalizing behavior ($\chi^2$ = 409.22, $p$ <.001, CFI = .88 and RMSEA = .07, 95% CI [.06, .08]) as well as internalizing behavior ($\chi^2$ = 443.80, $p$ <.001, CFI = .87 and RMSEA = .07, 95% CI [.07, .08]) had acceptable overall fit across multiple imputed data. The models show a significant association of child neglect and abuse with the intercepts of both internalizing and externalizing behaviors, however there was no association with status or development of dACC-anterior insula functional connectivity.
