## Supplement TableS1 for "Developing Functional Network Connectivity of the Dorsal Anterior Cingulate Cortex Mediates Externalizing Psychopathology in Adolescents with Child Neglect"

**Table S1.** Paths of parallel process latent growth models to link functional connectivity between dorsal anterior cingulate cortex and bilateral anterior insula and (1) externalizing and (2) internalizing behaviors.

| | $\beta$,*p* | 95% CI lower CI | 95% CI upper CI |
|---|---|---|---|
| **Model 1: Externalizing behavior** | | | |
| Neglect → $FC_{Intercept}$ | -.05 | -.18 | .08 |
| Neglect → $FC_{Slope}$ | .02 | -.11 | .20 |
| Neglect → $E_{Intercept}$ | .12* | .00 | .24 |
| Neglect → $E_{Slope}$ | .08 | -.08 | .23 |
| Abuse → $FC_{Intercept}$ | -.03 | -.14 | .11 |
| Abuse → $FC_{Slope}$ | -.04 | -.18 | .12 |
| Abuse → $E_{Intercept}$ | .15* | .03 | .29 |
| Abuse → $E_{Slope}$ | .00 | -.13 | .14 |
| $FC_{Intercept}$ → $E_{Slope}$ | -.06 | -.20 | .10 |
| $E_{Intercept}$ → $FC_{Slope}$ | .06 | -.06 | .19 |
| Neglect → $E_{Intercept}$ → $FC_{Slope}$ | .01 | -.00 | .04 |
| Neglect → $FC_{Intercept}$ → $E_{Slope}$ | .00 | -.01 | .02 |
| Abuse → $E_{Intercept}$ → $FC_{Slope}$ | .01 | -.01 | .04 |
| Abuse → $FC_{Intercept}$ → $E_{Slope}$ | .00 | -.01 | .02 |
| **Model 2: Internalizing behavior** | | | |
| Neglect → $FC_{Intercept}$ | -.06 | -.19 | .08 |
| Neglect → $FC_{Slope}$ | .00 | -.13 | .18 |
| Neglect → $I_{Intercept}$ | .13* | .00 | .24 |
| Neglect → $I_{Slope}$ | .12 | -.02 | .26 |
| Abuse → $FC_{Intercept}$ | -.01 | -.14 | .11 |
| Abuse → $FC_{Slope}$ | -.01 | -.16 | .13 |
| Abuse → $I_{Intercept}$ | .14* | .05 | .32 |
| Abuse → $I_{Slope}$ | -.00 | -.16 | .15 |
| $FC_{Intercept}$ → $I_{Slope}$ | .01 | -.16 | .18 |
| $I_{Intercept}$ → $FC_{Slope}$ | .01 | -.12 | .14 |
| Neglect → $I_{Intercept}$ → $FC_{Slope}$ | .01 | -.00 | .04 |
| Neglect → $FC_{Intercept}$ → $I_{Slope}$ | .00 | -.01 | .02 |
| Abuse → $I_{Intercept}$ → $FC_{Slope}$ | .01 | -.01 | .04 |
| Abuse → $FC_{Intercept}$ → $I_{Slope}$ | .00 | -.01 | .02 |

*Note.* FC = functional connectivity from dorsal anterior cingulate cortex to regions of the cingulo-opercular network, E = externalizing, I = internalizing, $\beta$ = regression weight, CI = confidence interval, *p*(*) = significant paths.
